# Reduced Cortical Excitability is Associated with Cognitive Symptoms in Concussed Adolescent Football Players

**DOI:** 10.1101/2024.09.23.24314232

**Authors:** Kevin C. Yu, Alex I. Wiesman, Elizabeth Davenport, Laura A. Flashman, Jillian Urban, Srikantan S. Nagarajan, Kiran Solingpuram Sai, Joel Stitzel, Joseph A. Maldjian, Christopher T. Whitlow

**Author notes:** Corresponding author, Kevin C. Yu and Alex I. Wiesman are equal contributors to this work and designated as co-first authors. <u>Correspondence</u>: Alex I. Wiesman, PhD, Department of Biomedical Physiology & Kinesiology Simon Fraser University, Burnaby, British Columbia, Christopher T. Whitlow, MD, PhD, MHA Department of Radiology, Wake Forest School of Medicine, Winston-Salem, North Carolina.

## Abstract

**Background:** American tackle football is associated with high rates of concussion, leading to neurophysiological disturbances and debilitating clinical symptoms. Previous investigations of the neurophysiological effects of concussion have largely ignored aperiodic neurophysiological activity, which is a marker of cortical excitability.

**Purpose:** We examined whether concussion during a season of high school football is related to changes in aperiodic and periodic neurophysiological activity and whether any such changes are associated with clinical outcomes.

**Materials and Methods:** Pre- and post-season resting-state magnetoencephalography (MEG) data were collected from 91 high school football players over as many as four seasons of play, for a total of 278 data collections. During these seasons of football play, a cohort of 10 individuals were diagnosed with concussion. MEG data were source-imaged, frequency-transformed and parameterized, and linear mixed models were used to examine effects of concussion on pre-to-post-season changes in neurophysiological activity. Scores on the Post-Concussive Symptom Inventory were correlated with pre-to-post-season neurophysiological changes to determine their clinical relevance.

**Results:** Concussion was associated with increased aperiodic exponents in superior frontal cortices, indicating a relative reduction in cortical excitability. This slowing of aperiodic neurophysiology mediated concussion effects on raw delta and gamma power and was associated with worse cognitive concerns across participants. Pre-to-post-season changes in aperiodic-corrected alpha and theta rhythmic activity were also decreased in posterior cortices in concussed players.

**Conclusion:** These findings indicate that concussion alters both the excitability and rhythmic signaling of the cortex, with differing spatial topographies and implications for clinical symptoms.

**Key Results:**

1. Concussion reduces cortical excitability in superior frontal cortices.
2. This reduction accounts for canonical effects of concussion on delta and gamma power.
3. Concussion-related changes in cortical excitability are associated with increased cognitive symptom severity.

## Introduction

Among youth sports played in the United States, tackle football is associated with the greatest number of concussions, with between 1.1 and 1.9 million detected annually in players 18 years old and younger.^1,2^ Concussions, also known as mild traumatic brain injuries (mTBI), cause axonal injury and disturbances in brain function, resulting in debilitating clinical sequelae both acutely and chronically. ^2–4^ The adolescent developing brain is especially susceptible to concussions, resulting in profound neurostructural and neurophysiological changes at the macro-scale. ^3,5–10^

Magnetoencephalography (MEG) can non-invasively detect these neurophysiological changes with millisecond time resolution,^11,12^ and has potential value as a clinical tool for concussion diagnosis and prognosis^5,13,14^. Previous examinations of mTBIs resulting from both football- and military-related impacts with MEG have shown abnormal changes in frequency-defined neurophysiological activity, manifesting most notably as alterations in delta (1 – 4 Hz) and gamma (30 – 80 Hz) activities, which are related to the severity of cognitive impairments.^5,10,13^ Concussion-related increases in slow-frequency activity in particular mirror similar pathological increases in patient with Parkinson’s^15–18^ and Alzheimer’s^19–24^ diseases, which are related to the hallmark proteinopathy and clinical impairments of each disorder.

With few exceptions^14,25^, previous M/EEG studies of concussion have used traditional approaches to spectral power analysis – exploring neurophysiological activity within canonical frequency bands while ignoring the aperiodic (i.e., arrhythmic) component. However, recent research has shown that aperiodic neurophysiological activity, and particularly its negative-signed slope (typically quantified as a positively-signed exponent), has substantial implications for understanding human brain function in states of health and disease ^18,26–36^. The aperiodic slope of the neural signal flattens with healthy aging^29,33^ and development^37–40^, and is slowed (i.e., steepens) in several neurological disorders such as Parkinson’s disease^18,41,42^, epilepsy^43,44^, and stroke^35^. Moreover, ignoring aperiodic activity as a source of “noise” runs the risk of conflating alterations in aperiodic neurophysiology with those in periodic neuronal oscillations^29,45^, thereby reducing sensitivity and interpretational clarity.

Investigations of the aperiodic slope in human neurophysiology have indicated its potential utility as a proxy of general cortical excitability. A previous EEG study of sleep and general anesthesia in adults demonstrated that the aperiodic slope can reliably track fast changes in cortical excitability across states of wakefulness, NREM, and REM sleep – specifically reporting steepening of the spectral slope during REM and propofol administration^27^, both of which are known to increase inhibition. Furthermore, an MEG study of generalized epilepsy showed that flattening of the aperiodic slope indexed whole brain hyperexcitability, even in the absence of interictal epileptic discharges^46^. These findings are also supported by computational modeling of the biophysical bases of human aperiodic neurophysiology^47^, and together indicate that steeper aperiodic slopes (i.e., higher exponents) are associated with decreased excitability and/or increased inhibition in cortical circuits.

One recent MEG study^14^ used spectral parameterization to determine the effect of mTBI on periodic neurophysiological activity. The authors found a reduction of beta-frequency periodic activity in participants with a mTBI but did not explicitly test whether aperiodic activity also differed significantly between the mTBI and control groups. An EEG study of adults with mTBI also examined parameterized power spectra, and found significant group differences in the peak frequency of alpha periodic activity within frontoparietal regions, but no differences in aperiodic neurophysiological activity.^25^ However, this study did not include participants in adolescence, which is a period of rapid neurodevelopmental change, especially within frontal lobe regions^48^ that are susceptible to head impacts.^49,50^ Further, neither of these studies measured neurophysiological activity prior to the mTBI event, potentially limiting their statistical sensitivity.

In this study, we examined whether the occurrence of a concussion during a season of football play is related to altered cortical excitability from pre- to post-season, as indexed by changes in aperiodic neurophysiological activity, and whether any such changes are associated with clinical outcomes. We hypothesized that concussion would be associated with pathologic slowing of aperiodic neurophysiologic activity, indexed by a more positive exponent and indicating a reduction in cortical excitability, and that these changes would relate to clinical symptomatology. We also expected that previously reported alterations in delta- and gamma-frequency neurophysiological activity in concussed individuals may actually be attributable to this aperiodic slowing effect. Finally, we anticipated to replicate periodic activity decreases in the alpha and beta frequency bands.

## Materials & Methods

### Participants

This study was reviewed and approved by the Wake Forest School of Medicine Institutional Review Board. Each participant and their parent provided assent and written informed consent, respectively, following detailed description of the study. All protocols complied with the Declaration of Helsinki. Exclusion criteria based on medical history questionnaire consisted of past neurological illness or major psychiatric disease, developmental disorders, medications that alter neurophysiology, concussion occurrence in the preceding year, or any contraindications to MRI or MEG scans (e.g., non-removable ferromagnetic implants).

High school football players in Winston-Salem, North Carolina were recruited as part of the imaging Telemetry and Kinematic modeLing (iTAKL) study^5,51^, which follows varsity and junior varsity football players throughout their season(s) of play to study the effects of impact exposure on the brain. Of the football players recruited from the 2012 to 2019 seasons, ninety-one players had complete and useable MEG data at both pre- and post-time points for at least one season (all male; mean age = 16.35), with 57 providing useable data for one season (mean age = 16.43), 22 for two seasons (mean age = 16.33), 10 for three seasons (mean age = 15.88), and two for four seasons (mean age = 16.75). This resulted in 139 seasons of repeated measures data across 91 individuals, for a total of 278 pre- and post-season neuroimaging data collection time points. Over the course of all seasons, 10 of these players were diagnosed with a concussion during the season of play (mean age = 15.99). Concussions were identified by a certified athletic trainer who attended all games and team practices. Players with possible concussions were referred to our collaborating concussion clinic for further evaluation and diagnosis by a Sports Medicine physician, where the Post Concussion Symptom Inventory (PCSI)^52^ was administered. The football players also underwent preseason and postseason T1 MRI scans for anatomical co-registration of their MEG data.

### MRI Acquisition and Processing

Participants underwent MRI scans on a 3T Siemens Skyra MRI scanner with a 32 channel, high-resolution head/neck coil (Siemens Medical, Malvern, PA). T1-weighted anatomical MRI were obtained via a 3D volumetric magnetization prepared rapid gradient echo (MPRAGE) sequence with 0.9 mm isotropic resolution (TR = 1900, TI = 900, TE = 2.93, FA = 9 degrees, 176 slices). Fiducial markers were placed on the nasion and bilateral preauricular regions prior to MRI for co-registration with participant MEG data. Participant anatomical MRIs were segmented and parcellated using *Freesurfer recon-all*^53^ with default settings.

### Magnetoencephalography Data Collection and Analyses

Eight minutes of eyes-open resting-state MEG data^54^ were collected using a 275-channel axial gradiometer CTF Omega 2005 system (VSM MedTech Ltd., Coquitlam, Canada) at a sampling rate of 1200 Hz with participants sitting in an upright position and fixating on a central point. Participants were monitored via real-time audio-video feeds during data acquisition. MEG data was processed using *Brainstorm*.^55^ The MEG data were resampled to 250 Hz, band passed filtered between 0.5-100 Hz, and notch filtered at 60 Hz (and harmonics). Bad data segments were identified and rejected manually. Independent component analysis (ICA) was used to detect and remove artifacts, including heartbeats, eyeblinks, and muscle artifacts based on their temporal and spatial topographies. The MEG data were coregistered with each subject’s individual structural MRI.^56^ The coregistered data were then source localized using a linearly constrained minimum variance (LCMV) beamformer implemented in *Brainstorm* with default settings, with source orientations unconstrained to the cortical surface. Welch’s method (window = 6s; 50% overlap) was used to transform the source-level time series data into the frequency domain. These data were parameterized using *specparam*^29^ (Brainstorm MATLAB version; frequency range = 1-40 Hz; Gaussian peak model; peak width limits = 0.5-12 Hz; maximum n peaks = 3; minimum peak height = 3 dB; proximity threshold = 2 standard deviations of the largest peak; fixed aperiodic; no guess weight). To represent the aperiodic components of the neurophysiological power spectrum, the offset and slope of the arrhythmic model fit were extracted per vertex. We also re-computed the parameterization of these spectra, excluding the lower frequencies (new frequency range = 10 – 40 Hz), to ensure that the aperiodic offset and/or legitimate periodic delta-frequency activity did not bias our estimation of the aperiodic exponent. The aperiodic spectra were then subtracted from the original Welch’s PSDs to derive the rhythmic (i.e., aperiodic-corrected) spectra.

The rhythmic and arrhythmic neurophysiological features were subsequently averaged over the vertices per each region of the Desikan-Killiany atlas^57^, and the rhythmic spectral data were averaged over canonical frequency bands (delta: 2 - 4 Hz, theta: 5 - 7 Hz, alpha: 8 - 12 Hz, beta: 15 - 29 Hz). This resulted in six maps of neurophysiological activity for each season and participant: the rhythmic activity in each of the four canonical frequency bands and the arrhythmic activity represented by the aperiodic exponent (i.e., slope) and offset. As indicated above, we also generated an additional map per each season and participant of the high-frequency aperiodic exponent. For each of these features, the pre-season brain maps were subtracted from the post-season maps to generate maps of change in neurophysiological activity per each season and participant. Band-averaged maps of the un-parameterized spectral data (delta: 2 - 4 Hz, theta: 5 - 7 Hz, alpha: 8 - 12 Hz, beta: 15 - 29 Hz, gamma: 30 – 59 Hz) were also generated using a similar approach, to determine the influence of parameterization on previously reported neurophysiological changes.

### Post-Concussive Symptom Inventory

The Post-Concussive Symptom Inventory (PCSI) is a 21-item questionnaire that is used to discriminate between athletes with and without concussion by quantifying symptom severity on a scale of 0-6. Parental PCSI data were collected for each participant with diagnosed concussions at time of evaluation. Items on the questionnaire are grouped into physical, emotional, and cognitive symptom clusters.^52,58^ We focused our hypotheses on the cognitive symptom cluster of the PCSI, which includes: feeling mentally “foggy”, difficulty concentrating, difficulty remembering, getting confused with directions or tasks, answering questions more slowly than usual, and feeling slowed down.^52,58^

### Statistics

A linear mixed-effects modeling approach was implemented to test the relationship between the occurrence of concussion and post-to-pre-season neurophysiological changes using the *lme4* package^59^ in *R*. Note that a major benefit of this approach is that it allows for the nesting of multiple repeated-measures (i.e., seasons) within each participant, while still utilizing all of the available data. Concussion occurrence was modeled as a categorical variable (i.e., concussion versus no-concussion in a given season), and age at pre-season scan, time between pre-season and post-season scans, and body mass index (BMI) were included as nuisance covariates. The final model took the following form:

> *neurophysiological change ∼ concussion + preseason age + time between scans + BMI + (1|participant)*

For each neurophysiological feature of interest, this model was fit per each region of the Desikan-Killiany atlas and the resulting p-values associated with the effect of concussion were corrected across regions via the Benjamini-Hochberg method^60^. Neurophysiological data were averaged over atlas regions that exhibited significant effects (with the same sign) for subsequent data visualization and mediation modeling (see below). Based on previous literature^5,10,61–64^ and our own hypotheses, we initially focused our hypotheses on the maps of delta (both corrected and uncorrected for the aperiodic spectra) and gamma (aperiodic-uncorrected only, as parameterization above ∼ 40 Hz is not advised^29^) activity, as well as the aperiodic features (i.e., slope and offset). All models were re-computed using an outlier exclusion threshold of normalized residuals ≥ the 97.5 percentile to confirm that our results were not influenced by outliers, and only results that survived this procedure are reported.

To examine associations between neurophysiological changes in concussion-sensitive brain regions and cognition, we averaged the post-to-pre-season changes in neurophysiological activity over brain regions where a significant effect of concussion was observed and correlated these values with the cognitive domain score of the PCSI. Non-parametric permutation testing was also used to partially account for the biases introduced by small sample sizes in cross-sectional correlation analyses using the *lmp* function in *R* (method = “Exact”).

Where appropriate, causal mediation analysis was used to test for indirect effects using a non-parametric bootstrapping approach with 10,000 simulations^65^ on region-averaged neurophysiological features.

## Results

### Aperiodic Neurophysiological Activity is Slowed by Concussion

We first examined the effect of concussion on changes in aperiodic neurophysiological activity from pre-to-post-season. The aperiodic exponent (region-wise *p*_FDR_’s < .05; region-average effect: *p* < .001, t = 4.80; Figure 2) and offset (region-wise *p*_FDR_’s < .05; region-average effect: *p* < .001, t = 5.04; Figure S1) were both increased from pre- to post-season in individuals who experienced a concussion, relative to those who did not. Frequency spectra averaged over a representative concussion-sensitive brain region (left caudal middle frontal) are shown in Figure S2 for the pre- and post-season timepoints for concussed and non-concussed individuals. These relationships remained intact when outliers were excluded (see *Methods: Statistics*), as did the effect on the aperiodic exponent when it was parameterized only using high-frequency neurophysiological activity (10 – 40 Hz; region-wise *p*_FDR_’s < .05; Figure S3). Pre-to-post-season changes in each neurophysiological feature for concussed and non-concussed participants can be seen in Figure S4.

**Figure 1.**
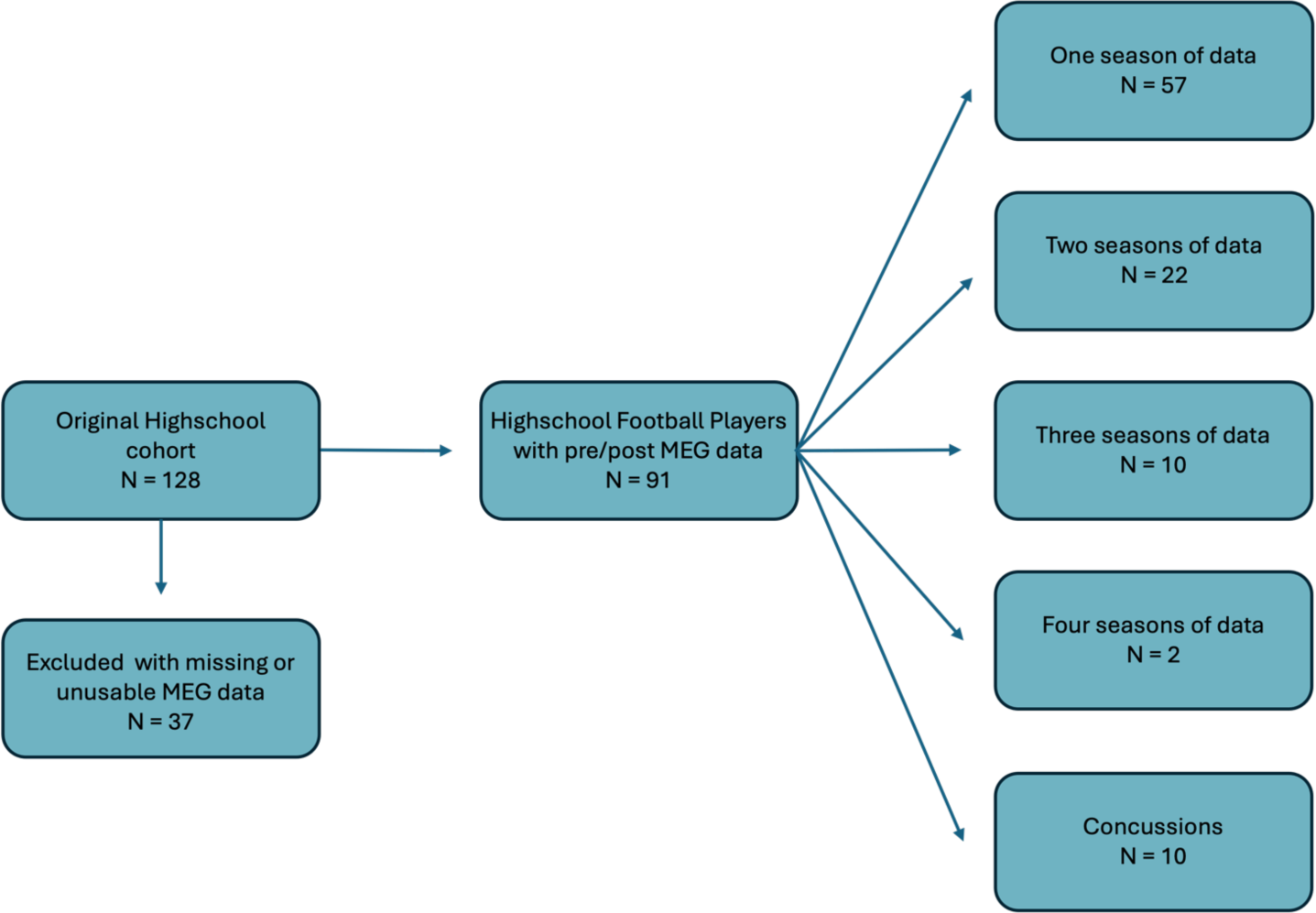
Participant Flowchart.

**Figure 2.**
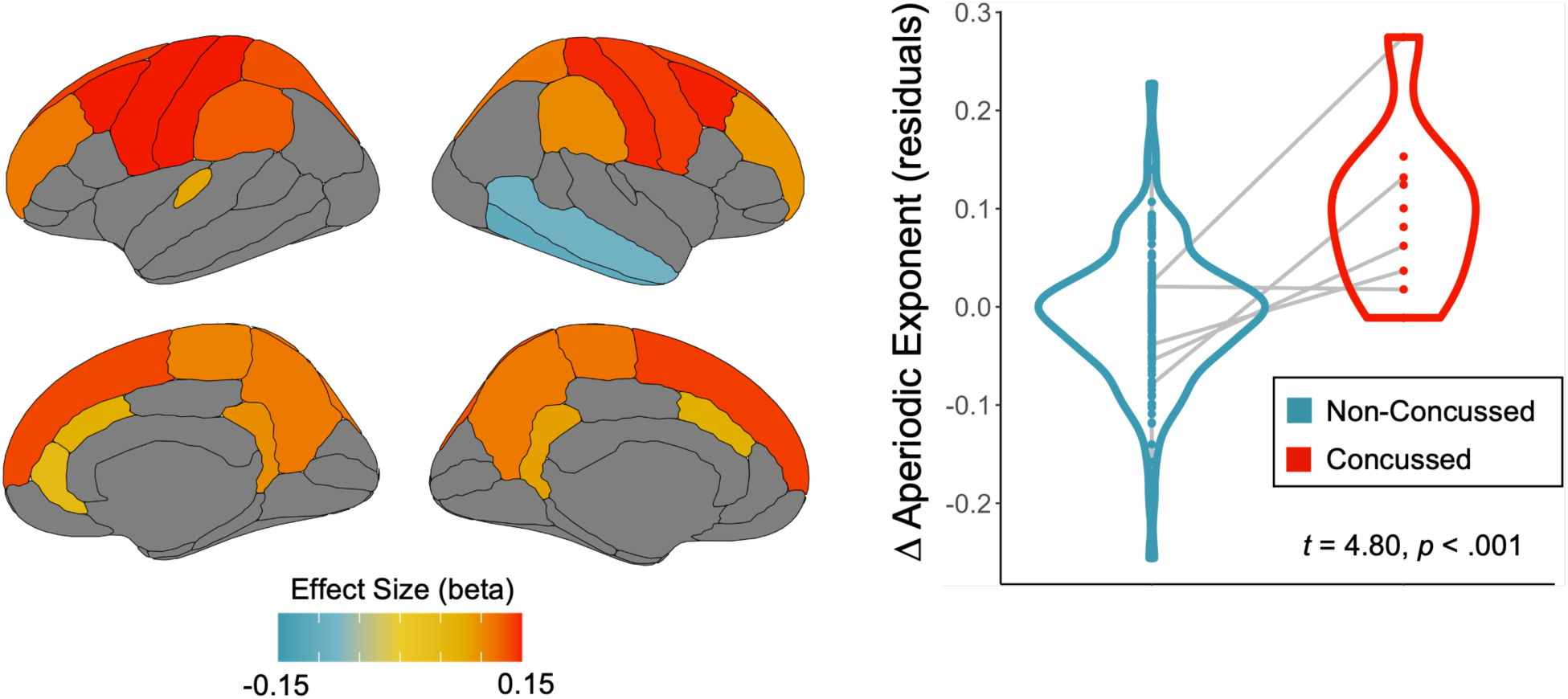
Concussion increases the aperiodic exponent in superior frontal cortices. Concussion effects on the post-minus-pre-season change in aperiodic exponent are displayed on the left as beta weight maps, thresholded based on a cutoff of *p*_FDR_ < .05. The violin plot to the right demonstrates the nature of this effect from values averaged over the warm-colored regions on the left, with the post-minus-pre-season change in aperiodic exponent (residuals) for concussed seasons in red and non-concussed seasons in blue.

### Aperiodic Slope Contributes Significantly to Delta and Gamma Power Abnormalities in Concussion

We replicated previously reported increases and decreases in raw delta and gamma power respectively in concussed participants (region-wise *p*_FDR_’s < .05; Figure 3), and the spatial topography of this effect qualitatively mirrored the concussion-related slowing of aperiodic neurophysiological activity. However, once we corrected the delta band values for the aperiodic features, we observed no such effect (all region-wise *p*_FDR_’s > .309; Figure 3). Note that similar correction for the aperiodic component in the gamma band was not possible, due to issues with fitting 1/f models at frequencies above ∼40 Hz.^29^ We also found that the observed change in the aperiodic exponent accounted for the effect of concussion on both delta and gamma power using causal mediation analysis. There was an indirect effect of the aperiodic exponent on both the concussion-raw delta relationship (ACME = .020, *p* < .001; Figure 3) and the concussion-raw gamma relationship (ACME = -.002, *p* < .001; Figure 3).

**Figure 3.**
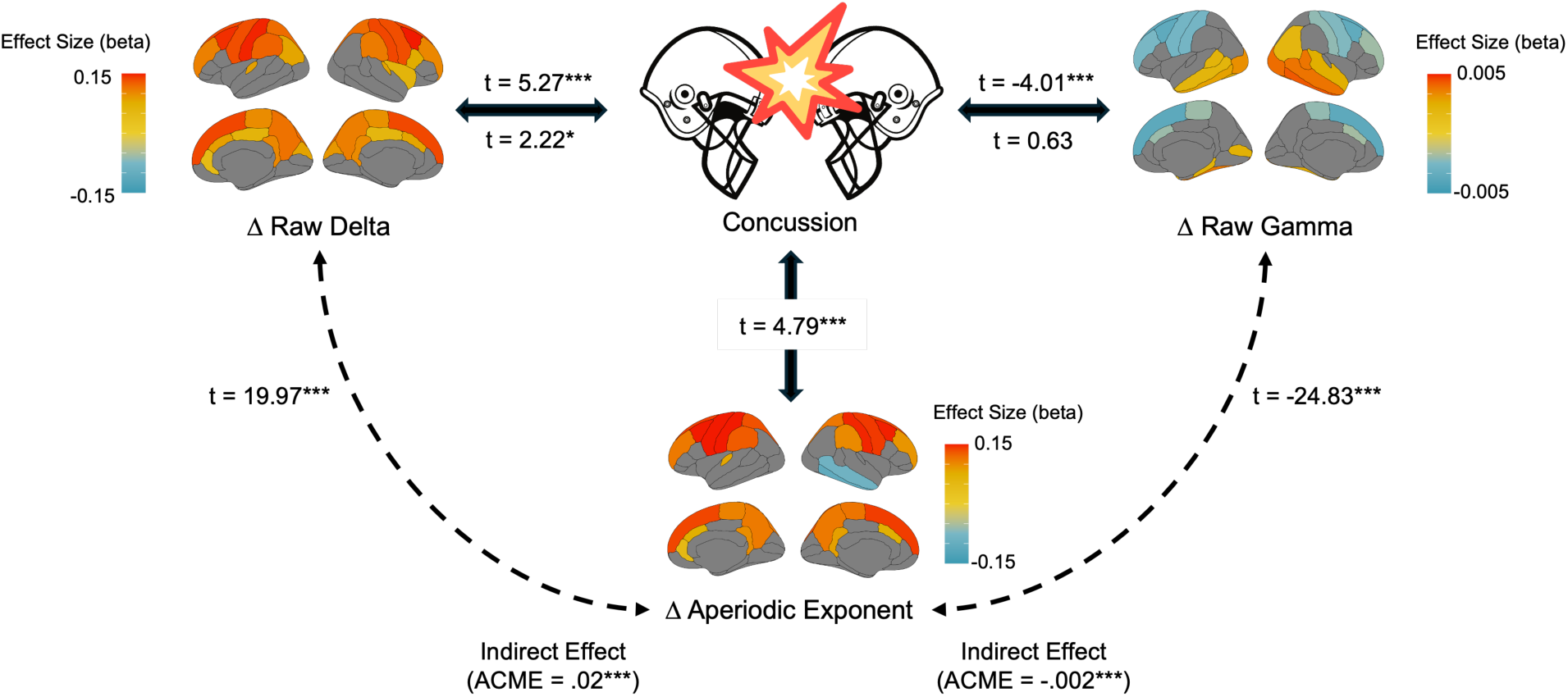
Aperiodic slowing accounts for the effect of concussion on raw delta and gamma power. Causal mediation analysis: regression paths are shown between concussion (top middle), post-minus-pre-season change in aperiodic exponent (bottom; averaged over significant regions from the linear-mixed model analysis in Figure 2), raw delta band power (top left; averaged over the same regions from Figure 2), and raw gamma band power (top right; averaged over the same regions from Figure 2). The t-values next to each line correspond to their linear relationship modeled via multiple regression (accounting for nuisance covariates). The t-values above the top left and right arrows represent the relationships between concussion and changes in raw delta and gamma activity, respectively, without accounting for changes in the aperiodic exponent. The t-values below the top left and right arrows indicate the same relationships once change in the aperiodic exponent was included in the model. The t-values outside the dotted lines represent the relationships between post-minus-pre-season changes in aperiodic exponent and changes in raw delta and gamma power. The average causal mediation effects (ACME; below) indicated significant mediation by the aperiodic exponent for both the concussion-delta and concussion-gamma relationship. \**p* < 0.05, \*\*\**p* < 0.001.

### Periodic Alpha and Theta Activity is Reduced by Concussion

We also examined whether aperiodic-corrected neurophysiological activity was related to the occurrence of concussion. Aperiodic-corrected alpha (region-wise *p*_FDR_’s < .05; region-average effect: *p* < .001, t = -4.57; Figure 4) and theta (region-wise *p*_FDR_’s < .05; region-average effect: *p* < .001, t = -4.17; Figure S5) activity were both decreased from pre- to post-season in individuals who experienced a concussion, relative to those who did not. These relationships remained intact when outliers were excluded (see *Methods: Statistics*). No significant effects were seen in the aperiodic-corrected delta and beta bands.

**Figure 4.**
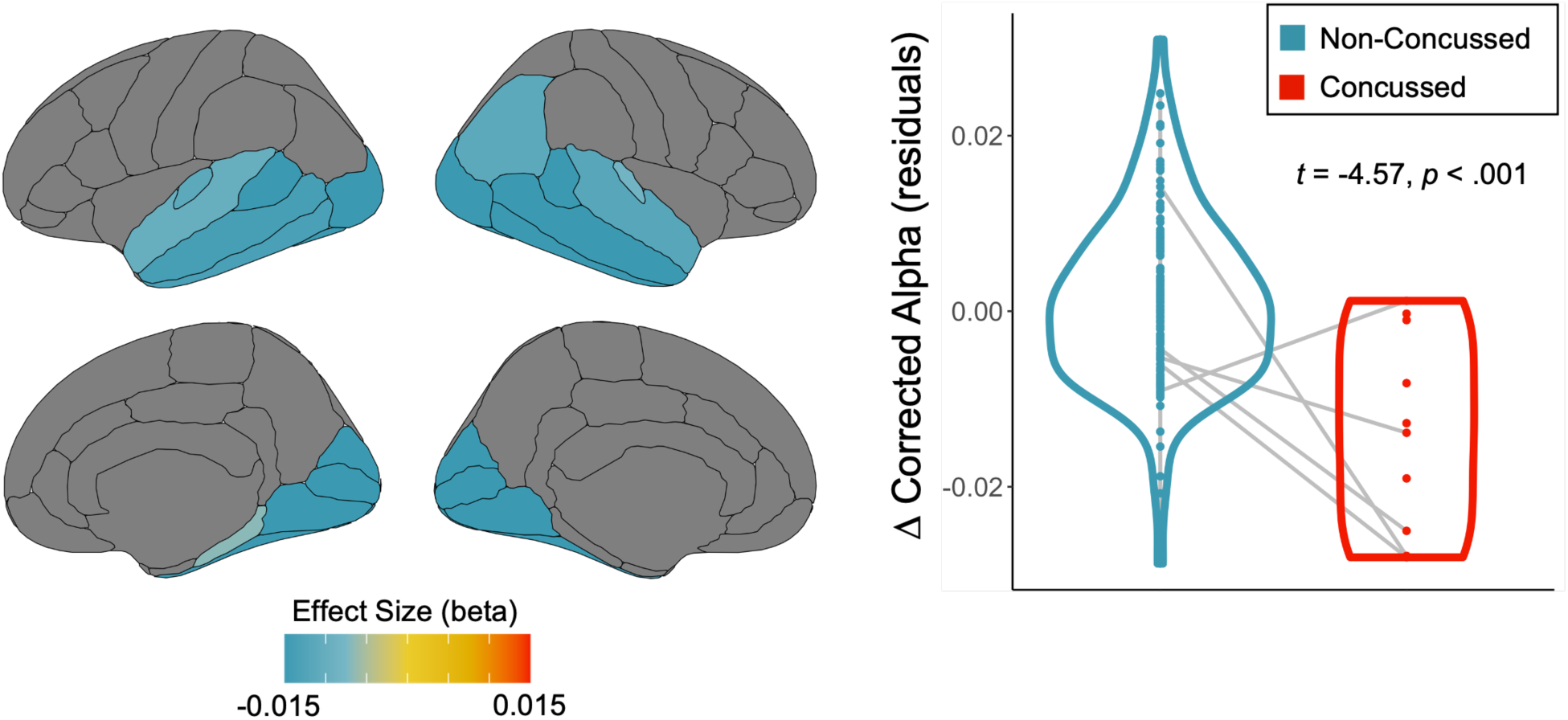
Concussion decreases aperiodic-corrected alpha oscillations in temporo-occipital cortices. Concussion effects on the post-minus-pre-season change in aperiodic corrected alpha band are displayed on the left as beta weight maps, thresholded based on a cutoff of *p*_FDR_ < .05. The violin plot to the right demonstrates the nature of this effect from values averaged over the filled regions on the left, with the post-minus-pre-season change in aperiodic exponent (residuals) for concussed seasons in red and non-concussed seasons in blue.

### Concussion-related Aperiodic Slowing is Associated with Severity of Cognitive Symptoms

Finally, to determine the clinical relevance of the observed concussion-related changes in the aperiodic exponent, we related PCSI cognitive symptom scores to changes in the pre-to-post-season exponents that we averaged over the concussion-sensitive regions indicated by warm colors in Figure 2. There was a significant positive correlation between the PCSI cognitive domain scores and pre-to-post-season exponent changes (*r* = .85, *p* = .008; N = 8; Figure 5), such that greater concussion-related aperiodic slowing was associated with higher reported cognitive symptoms at the time of assessment. To partially address the limitations of the small sample size, we re-computed this relationship using nonparametric permutation testing and findings remained significant (10,000 permutations; *p*_PERM_ = .012).

**Figure 5.**
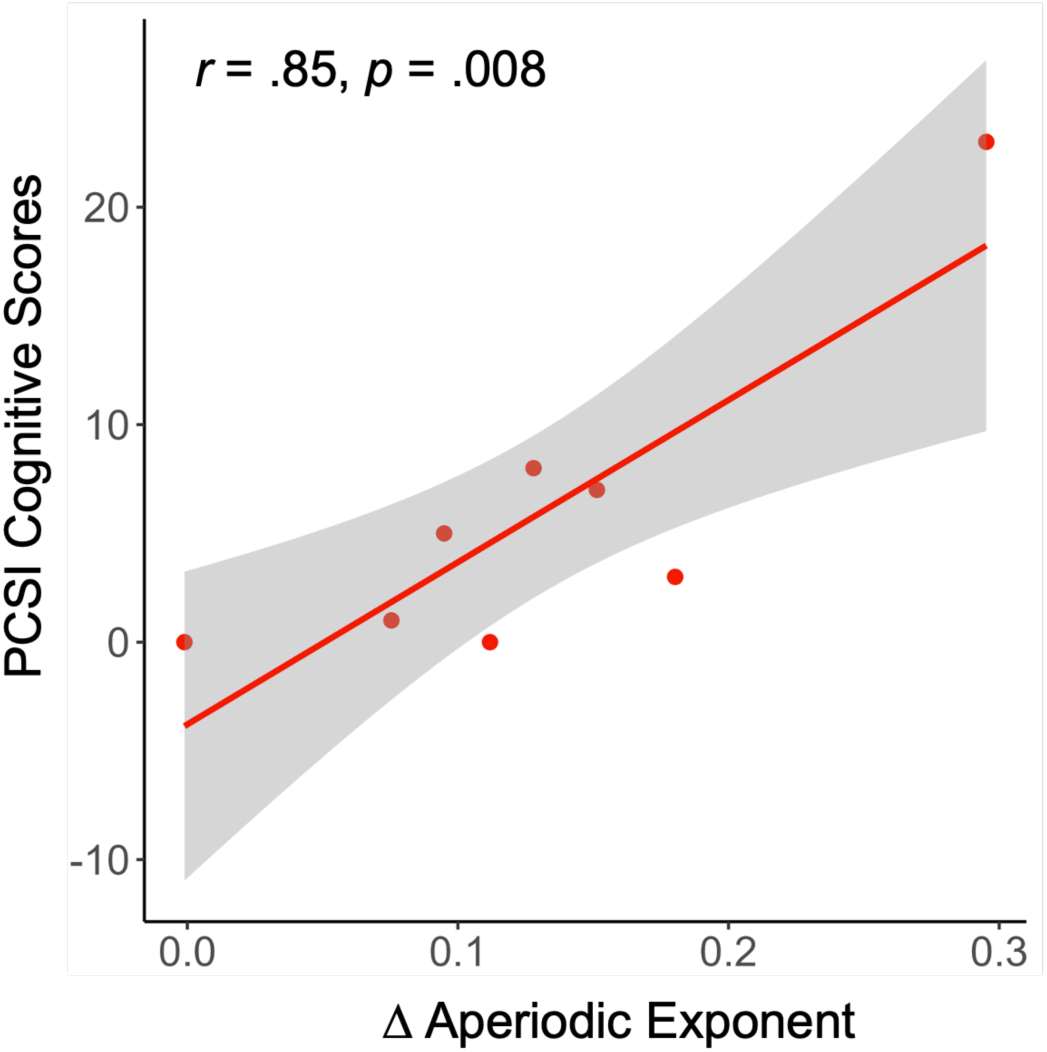
Concussion-related changes in aperiodic exponent are associated with the severity of cognitive symptoms. Post-minus-pre-season changes in the aperiodic exponent were averaged over the warm-colored concussion-sensitive brain regions indicated in Figure 2 and are plotted (x-axis) against the cognitive symptom scores from the Post-Concussion Symptom Inventory (PCSI; y-axis). The line-of-best fit and associated statistics are overlaid, and the shaded area indicates the 95% confidence interval. Note that of the 10 participants with complete MEG data who experienced a concussion, only N = 8 also had PCSI scores available for this analysis.

## Discussion

Alterations in neurophysiological activity have been previously documented in adults with mTBI^13^ and to a lesser degree, in pediatric populations^5^; however, these investigations have primarily focused on periodic (i.e., rhythmic) brain activity. Whether the emerging physiologic importance of the aperiodic component of neurophysiological activity is related to concussion in the adolescent brain, and whether any such changes are clinically meaningful, are questions that have yet to be answered. Using resting-state MEG, we demonstrate aperiodic neurophysiologic slowing after concussion in youth tackle football players, indicating reductions in cortical excitability, with association to cognitive symptoms.

With these same data, and in agreement with several reports^10,66–70^, Davenport et al. previously showed that whole-brain delta power was significantly increased in high school American football players after a concussion was diagnosed during the season.^5^ We reaffirm this finding with enhanced spatial specificity: activity in the delta frequency band, when uncorrected for aperiodic activity, is increased from pre- to post-season in adolescents who experienced concussion in this participant sample. We also extend our analyses to the gamma band and find a significant decrease in gamma activity after concussion, which again aligns with previous research.^13,71^

Intriguingly, we then showed that the aperiodic exponent increased from pre- to post-season in tackle football players with diagnosed concussion, relative to players without such a diagnosis, indicating a relative decrease in cortical excitability. This aligns with previous research indicating cortical hypoexcitability and increased inhibition following concussion^72,73^. The spatial topography of this concussion effect is similar to those of raw delta and gamma activity, indicating that it may be responsible for the effects seen in these canonical frequency bands. Supporting this concept, we find that correction of delta band values for aperiodic activity results in no observed concussion-related effects in this band. Additionally, a causal mediation analysis indicated that the concussion-raw delta and concussion-raw gamma relationships could both be explained by changes in the aperiodic exponent. Using the parental PCSI, which has high test-retest reliability and developmentally appropriate utility in studying concussion-related symptomatology,^58^ we also show that the magnitude of concussion-related decreases in cortical excitability are associated with higher parent report of cognitive symptoms in concussed adolescents, indicating potential for the use of aperiodic features as clinical markers.

We also observe significant pre- to post-season decreases in both alpha and theta periodic activity in posterior cortical regions, which replicate and add to existing literature indicating rhythmic changes in the context of mTBI.^14,25^ We did not observe the decrease in beta band activity that has previously been reported in adults with mTBI, and future research should investigate potential developmental changes in the spectral definitions of periodic effects of mTBI.

Our results are notably limited by a relatively small sample of individuals who experienced a concussion during a season of youth football, due to the inherent difficulties of collecting this type of multi-timepoint data. However, this repeated measures approach (i.e., both from pre-to-post season and across seasons within an individual) partially ameliorates this concern by reducing unrelated variance in our signals of interest, and our linear mixed-effects statistical approach fully leverages the nested nature of these data. The inclusion of only male participants is also a limitation and should be addressed via similar studies in female and mixed-sex impact sports. The subjectivity regarding the clinical diagnosis of concussion is also important to consider in this study; however, several factors were employed to mitigate this limitation. Not only were parents, coaches, and players educated before each season on the signs and symptoms of concussion, but an experienced certified athletic trainer was also present at all practices and games to help identify these signs and assess for concussion. Furthermore, a board-certified sports medicine physician evaluated all suspected concussions at our collaborating concussion clinic within 24-72 hours of injury to confirm the diagnosis.

Overall, our findings suggest that previous electrophysiological neuroimaging findings in mTBI could be, at least partially, explained by a more parsimonious slowing of aperiodic neurophysiological activity, which is also associated with symptom severity. Our results in parallel demonstrate the utility of parameterizing the neurophysiological signal to enhance mechanistic interpretation and statistical sensitivity.

**Table 1.** Clinical Characteristics of Participants.

|  | Concussion (N = 10) | Without Concussion (N = 81) |
| --- | --- | --- |
| **Demographics** |  |  |
| Age (years) | 15.99±.85 | 16.13±1.19 |
| BMI | 25.87±6.69 | 26.75±5.41 |
| Education completed (years) | 9.22±.83 | 9.57±1.09 |
| Time Between Scans (days) | 143.2±23.33 | 159.04±49.63 |
| Handedness (% Right Handed)* | 9(100) | 54(87) |
| **Race or ethnicity** |  |  |
| Native American | 0 | 3(3.7) |
| Asian | 0 | 1(1.23) |
| Pacific Islander | 0 | 1(1.23) |
| Black | 5(50) | 33(40.74) |
| White | 5(50) | 44(54.32) |
| Hispanic | 0 | 1(1.23) |
| Non-Hispanic | 8(80) | 43(53.09) |

Values for continuous measures are means±1 standard deviation.
Categorical variables are shown as numbers of participants where data was available, with percentages in parentheses.
\*Handedness data was only provided by nine of the concussed participants and 62 of 81 football players without concussion.

## Data Availability

All data produced in the present study are available upon reasonable request to the authors

## Acknowledgments

This work was supported by the National Institutes of Health (NIH) grants R01NS082453 and R01NS091602 to CTW; a Banting Postdoctoral Fellowship (BPF-186555) and the Canada Research Chair (CRC-2023-00300) in Neurophysiology of Aging and Neurodegeneration from the Canadian Institutes of Health Research (CIHR) and NIH grant F32-NS119375 to AIW. Additionally, special thanks to Wake Forest School of Medicine and the Childress Institute for Pediatric Trauma at Wake Forest Baptist Medical Center for the support provided during the study.

