## Supplement for "Reduced Cortical Excitability is Associated with Cognitive Symptoms in Concussed Adolescent Football Players"

**Supplementary Materials**

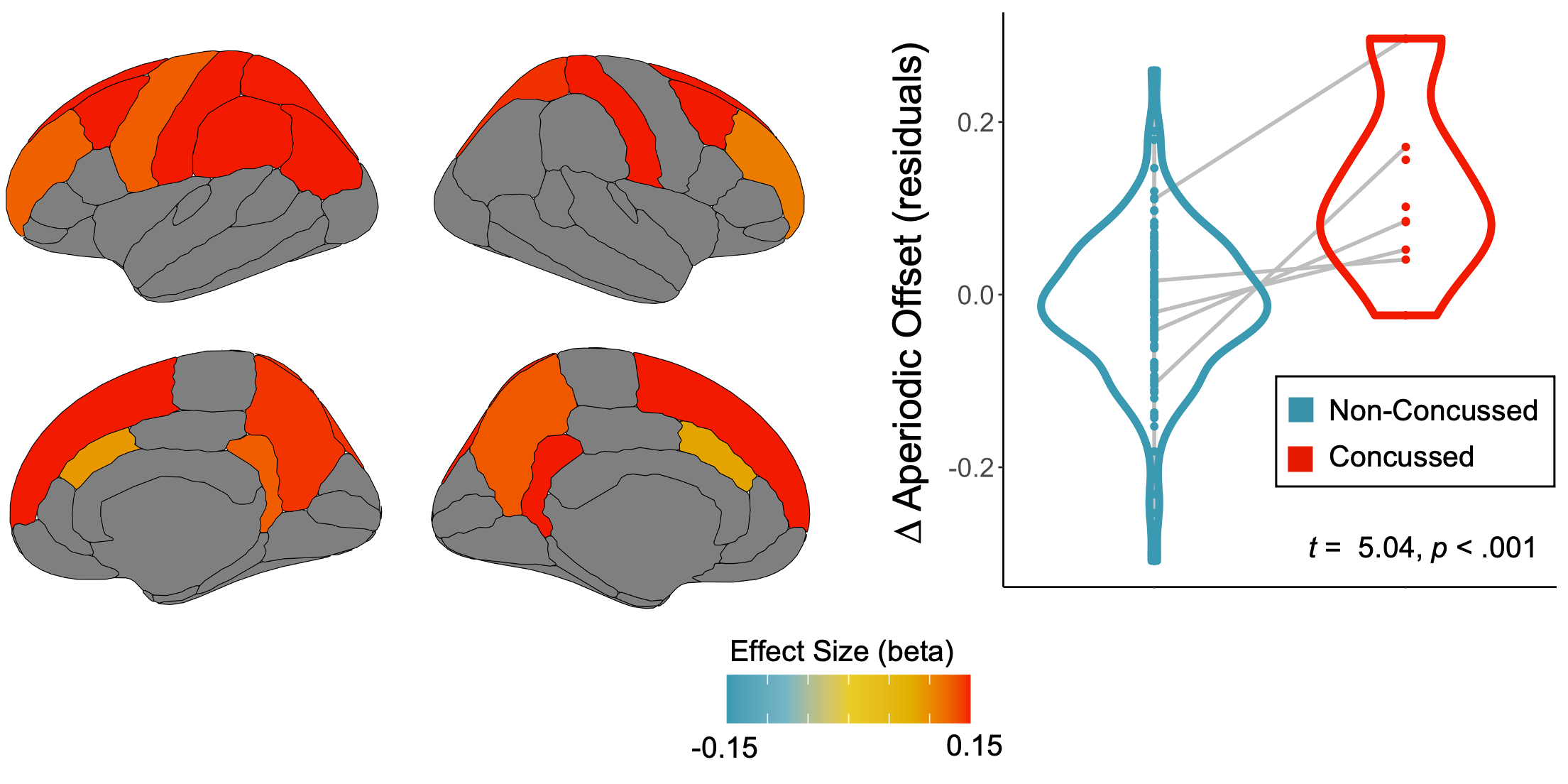
**Figure S1. Concussion increases the aperiodic offset in superior frontal cortices.** Similar to Figure 2, but for the aperiodic offset rather than the aperiodic exponent.

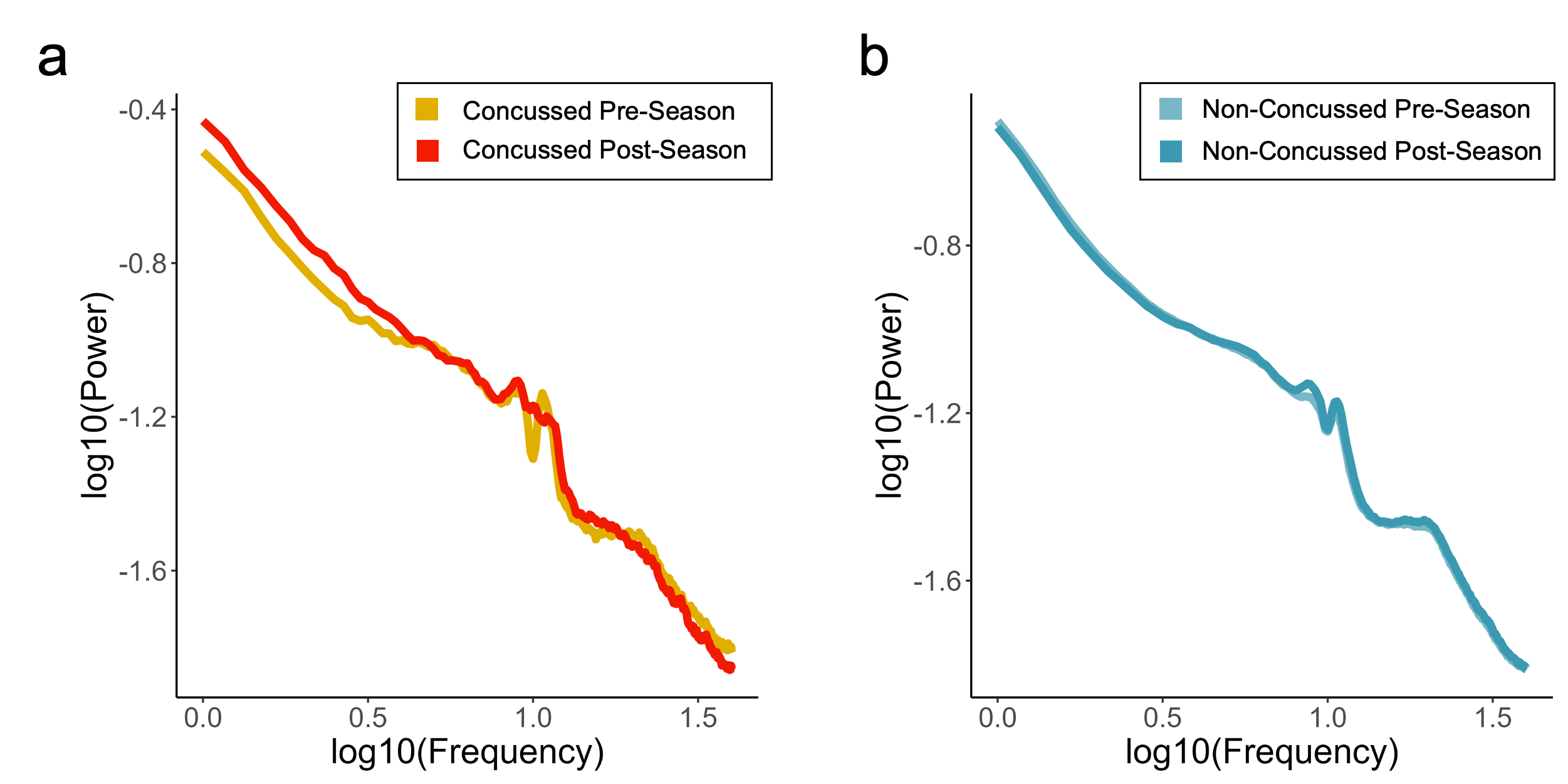

**Figure S2. Averaged frequency spectra for pre- and post-seasons for concussed and non-concussed individuals in left caudal middle frontal region.** Participant spectra were averaged within each of the indicated sub-groups over the left caudal middle frontal region, which exhibited the strongest effect shown in Figure 2.

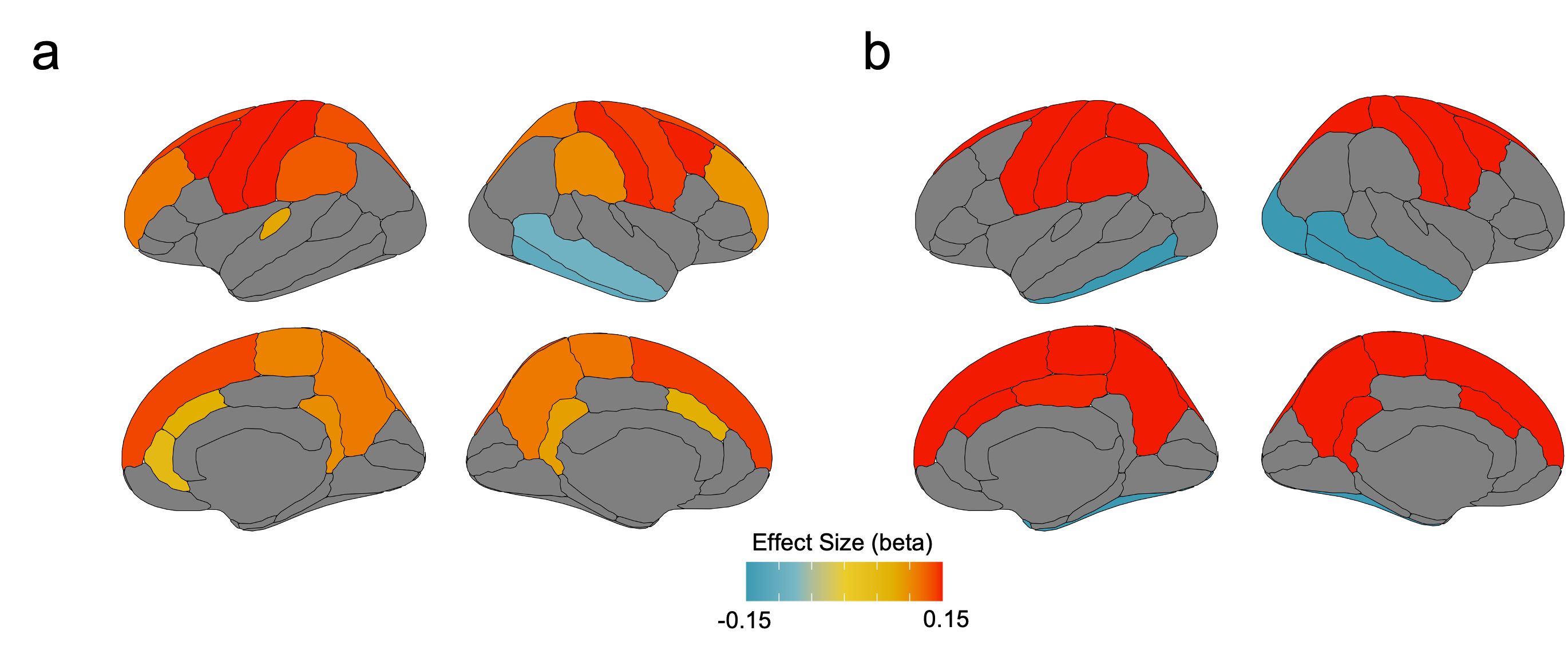
**Figure S3. Concussion increases the aperiodic exponent in superior frontal cortices (parameterized using both 1 – 40Hz and 10 – 40Hz frequency ranges).** Similar to Figure 2, with parameterization of the aperiodic exponent performed using (a) the original 1 – 40Hz default range and (b) a higher 10 – 40Hz frequency range which excludes low-frequency/delta activity.

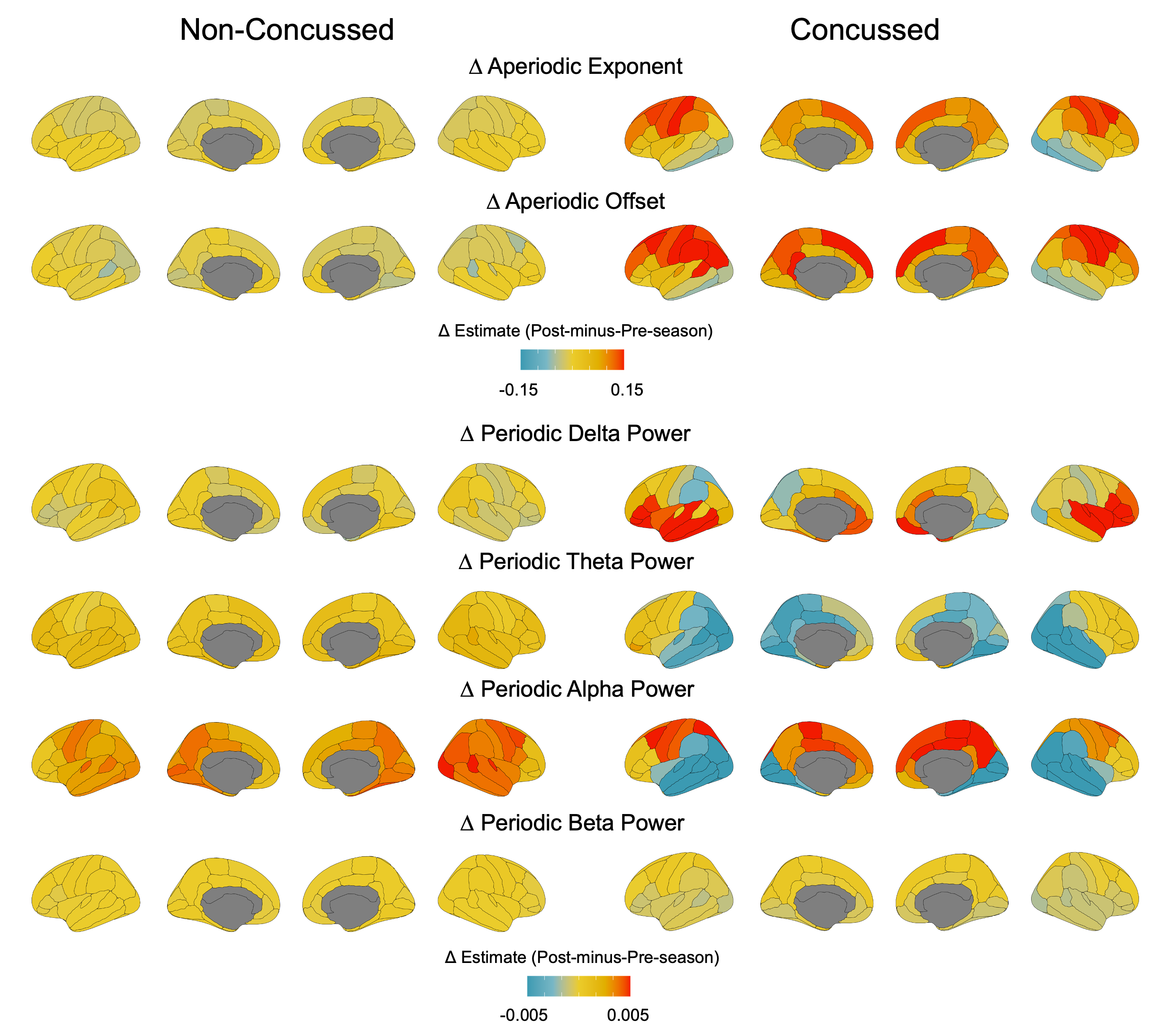

**Figure S4. Post-to-pre-season changes in aperiodic and periodic neurophysiological activity for concussed and non-concussed participants.**

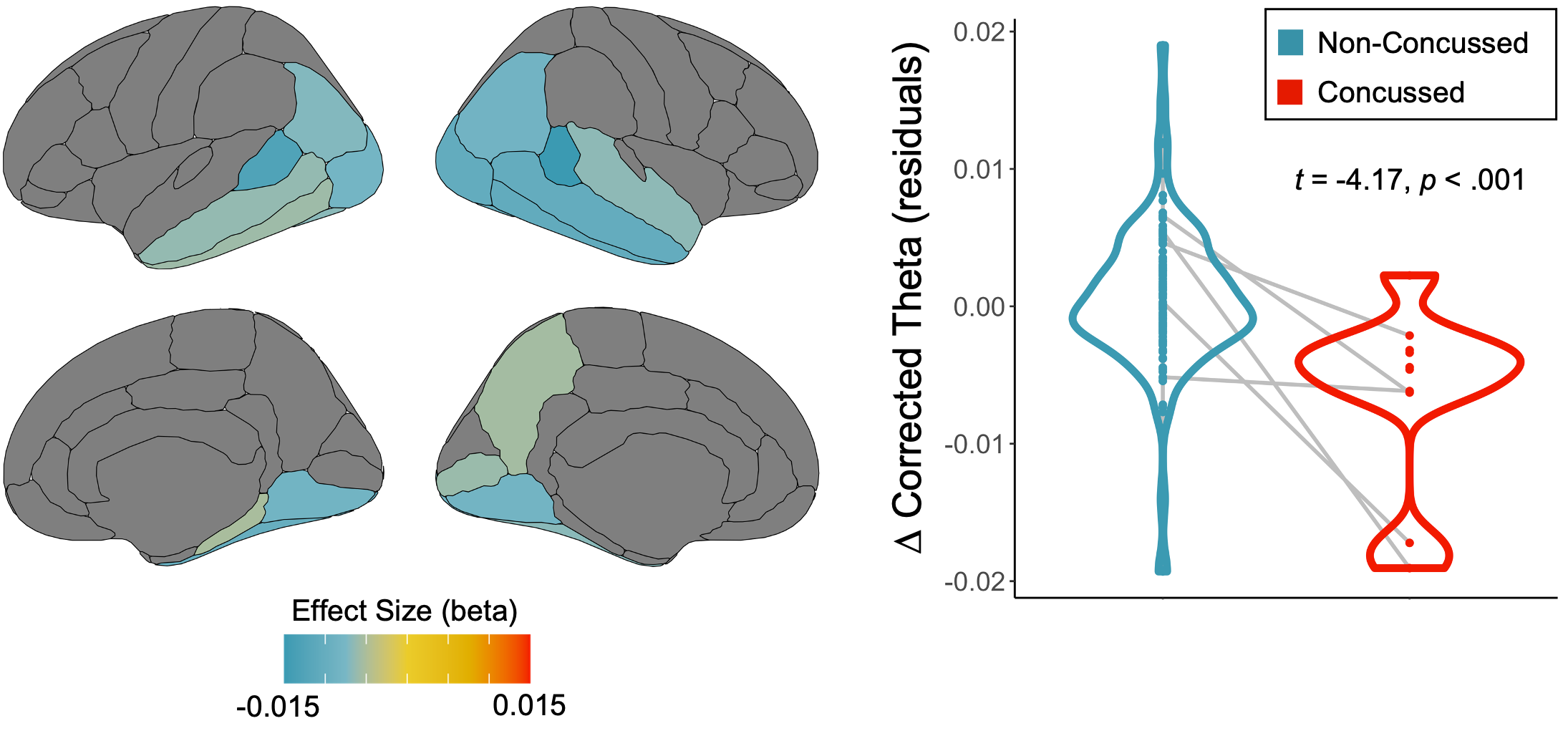

**Figure S5. Concussion decreases aperiodic-corrected theta oscillations in temporo-occipital cortices.** Concussion effects on the post-minus-pre-season change in aperiodic-corrected theta band activity are displayed on the left as beta weight maps, thresholded at *p*_FDR_ < .05. The violin plot to the right demonstrates the nature of this effect from values averaged over the filled regions on the left, with the post-minus-pre-season change in aperiodic exponent (residuals) for concussed seasons in red and non-concussed seasons in blue.

| **Desikan-Killiany Atlas Region** | ***p*_FDR_** | **effect (beta)** |
| --- | --- | --- |
| Caudal anterior cingulate L | 0.006 | 0.07 |
| Caudal anterior cingulate R | 0.006 | 0.07 |
| Caudal middle frontal L | <.001 | 0.15 |
| Caudal middle frontal R | 0.005 | 0.15 |
| Isthmus cingulate L | 0.016 | 0.09 |
| Isthmus cingulate R | 0.009 | 0.10 |
| Paracentral L | 0.005 | 0.11 |
| Paracentral R | 0.004 | 0.11 |
| Postcentral L | <.001 | 0.17 |
| Postcentral R | <.001 | 0.15 |
| Precentral L | <.001 | 0.15 |
| Precentral R | <.001 | 0.14 |
| Precuneus L | 0.002 | 0.11 |
| Precuneus R | 0.004 | 0.11 |
| Rostral anterior cingulate R | 0.042 | 0.05 |
| Rostral middle frontal L | <.001 | 0.11 |
| Rostral middle frontal R | 0.005 | 0.09 |
| Superior frontal L | <.001 | 0.14 |
| Superior frontal R | <.001 | 0.14 |
| Superior parietal L | 0.003 | 0.13 |
| Superior parietal R | 0.009 | 0.11 |
| Supramarginal L | 0.005 | 0.13 |
| Supramarginal R | 0.026 | 0.10 |
| Transverse temporal L | 0.024 | 0.08 |
| Inferior temporal R | 0.014 | -0.11 |
| Middle temporal R | 0.038 | -0.09 |

**Table S1. Region-wise beta weights and p-values for each cortical region from Figure 2 exhibiting a significant effect of concussion on the aperiodic exponent.**
